# Evaluating the Diagnostic and Treatment Recommendation Capabilities of GPT-4 Vision in Dermatology

**DOI:** 10.1101/2024.01.24.24301743

**Authors:** Abhinav Pillai, Sharon Parappally Joseph, Jason Kreutz, Danya Trabousi, Maharshi Gandhi, Jori Hardin

## Abstract

**Background:** The integration of artificial intelligence (AI) in dermatology presents a promising frontier for enhancing diagnostic accuracy and treatment planning. However, general purpose AI models require rigorous evaluation before being applied to real-world medical cases.

**Objective:** This project specifically evaluates GPT-4V’s performance in accurately diagnosing and generating treatment plans for common dermatological conditions, comparing its assessment of textual versus image data and its performance with multimodal inputs. Beyond the immediate scope, this study contributes to the broader trajectory of integrating AI in healthcare, highlighting the limitations of these technologies, as well as their potential to enhance efficiency, and education within medical training and practice.

**Methods:** A dataset of 102 images representing nine common dermatological conditions was compiled from open-access websites. Fifty-four images were ultimately selected by two board-certified dermatologists as being representative and typical of the common conditions. Additionally, nine clinical scenarios corresponding to these conditions were developed. GPT-4V’s diagnostic capabilities were assessed in three setups: Image Prompt (image-based), Scenario Prompt (text-based), and Image and Scenario Prompt (combining both modalities). The model’s performance was evaluated based on diagnostic accuracy, differential diagnosis, and treatment recommendations.

**Results:** In the Image Prompt setup, GPT-4V correctly identified the primary diagnosis for 29 of 54 images. The Scenario Prompt setup showed a higher accuracy rate of 89% in identifying the primary diagnosis. The multimodal Image and Scenario Prompt setup also achieved an 89% accuracy rate. However, a notable bias towards textual data over visual data was observed. Treatment recommendations were evaluated by the same two dermatologists, using a modified Entrustment Scale, showing competent but not expert-level performance.

**Conclusion:** GPT-4V demonstrates promising capabilities in dermatological diagnosis and treatment recommendations, particularly in text-based scenarios. However, its performance in image-based diagnosis and integration of multimodal data highlights areas for improvement. The study underscores the potential of AI in augmenting dermatological practice, emphasizing the need for further development, and fine-tuning of such models to ensure their efficacy and reliability in clinical settings.

## Introduction

### Background

The flourishing domain of Artificial Intelligence (AI) in healthcare, notably Generative Pre-trained Transformer models like GPT-4, heralds a promising era, especially with the recent advent of GPT-4 Vision (GPT-4V), a state-of-the-art, multimodal large language model (LLM) capable of processing both image and text inputs^1^. This development is of particular significance in dermatology, a field inherently reliant on visual data for accurate diagnostics and treatment planning. Past evaluations of GPT models using United States Medical Licensing Examination (USMLE) questions in dermatology were somewhat constrained as they could not utilize the accompanying images due to the lack of vision capabilities in earlier GPT models ^2,3^. However, with the introduction of GPT-4V, there’s potential to overcome this limitation, opening new avenues for more accurate and comprehensive dermatological assessments^1^.

Recent literature underscores the strides made with GPT models in dermatology^4,5^. Kluger highlighted the potential applications of ChatGPT in dermatology, especially in accurate disease identification and differential diagnosis^5^. Moreover, a systematic review underscored the proficiency of ChatGPT in dermatology, particularly in the domain of cancer, albeit with room for improvement in certain specialized areas like triaging the appropriate use of Mohs surgery for cutaneous neoplasms^6^. While ChatGPT has demonstrated some proficiency in handling patient queries in dermatology, the recent development of GPT-4V, with its ability to process both text and image inputs, presents an even greater opportunity for LLMs to augment dermatological assessments^7^. However, there is a need to evaluate the capabilities and limitations of GPT-4V in image and text based dermatological cases.

### Objective

This project evaluates the performance of GPT-4V’s capabilities at accurately diagnosing and formulating treatment plans for common dermatological conditions and compares the model’s performance in assessing textual versus image data and its performance with multimodal inputs, combining text and image data. Additionally, this work evaluates the broader trajectory of integrating AI in healthcare to foster a more efficient, educated, and error-minimized medical paradigm, thereby contributing to the existing body of knowledge regarding the application of AI in healthcare and medical education.

## Methods

### Image Collection

To address privacy and copyright challenges associated with utilizing images from established medical examinations or proprietary sources, our project compiled a dataset of 102 images with predetermined diagnoses from publicly available sources, specifically dermnet.nz and dermatlas.org. The chosen images represented 9 common dermatological conditions and were obtained from open-access platforms to ensure compliance with patient privacy regulations in Canada while still contributing to a robust evaluation of GPT-4V in dermatological applications.

The accuracy and quality of the images were evaluated by two board-certified dermatologists (JH and DT). While acknowledging that an individual image might exhibit features of various dermatological conditions, the stringent selection criteria required that included images showcase classic manifestations of their respective conditions, with the predominant representation being the condition of interest. Inclusion in the final analysis required the unanimous agreement of both reviewers. Using this inclusion criteria, 54 images were selected for the final analysis, with 6 images representing each of the 9 dermatological conditions.

### Clinical Scenario Creation

To provide a realistic assessment of GPT-4’s capabilities in dermatological diagnosis and treatment planning, a set of 9 unique clinical scenarios were created, specifically geared towards the medical student level. Each scenario corresponded to one of the 9 dermatological conditions under investigation. These scenarios represented the typical presentations of these conditions, encompassing common symptoms, patient history, and visual indicators ordinarily encountered in clinical practice (Supplementary Table 1).

To ensure the accuracy and relevance of these scenarios, they were reviewed and approved by two board-certified dermatologists (JH and DT).

### Diagnostic Evaluation

The diagnostic evaluation of GPT-4V was conducted in three distinct setups utilizing the ChatGPT platform, specifically, the version released on September 25, 2023. In each setup, a different type of prompt was used to assess the model’s ability to provide accurate diagnoses and relevant treatment recommendations based on the input data (Table 1). The prompts were constructed to reflect common clinical inquiries and were inputted into GPT-4V via the ChatGPT interface (Figure 1).

**Table 1:**
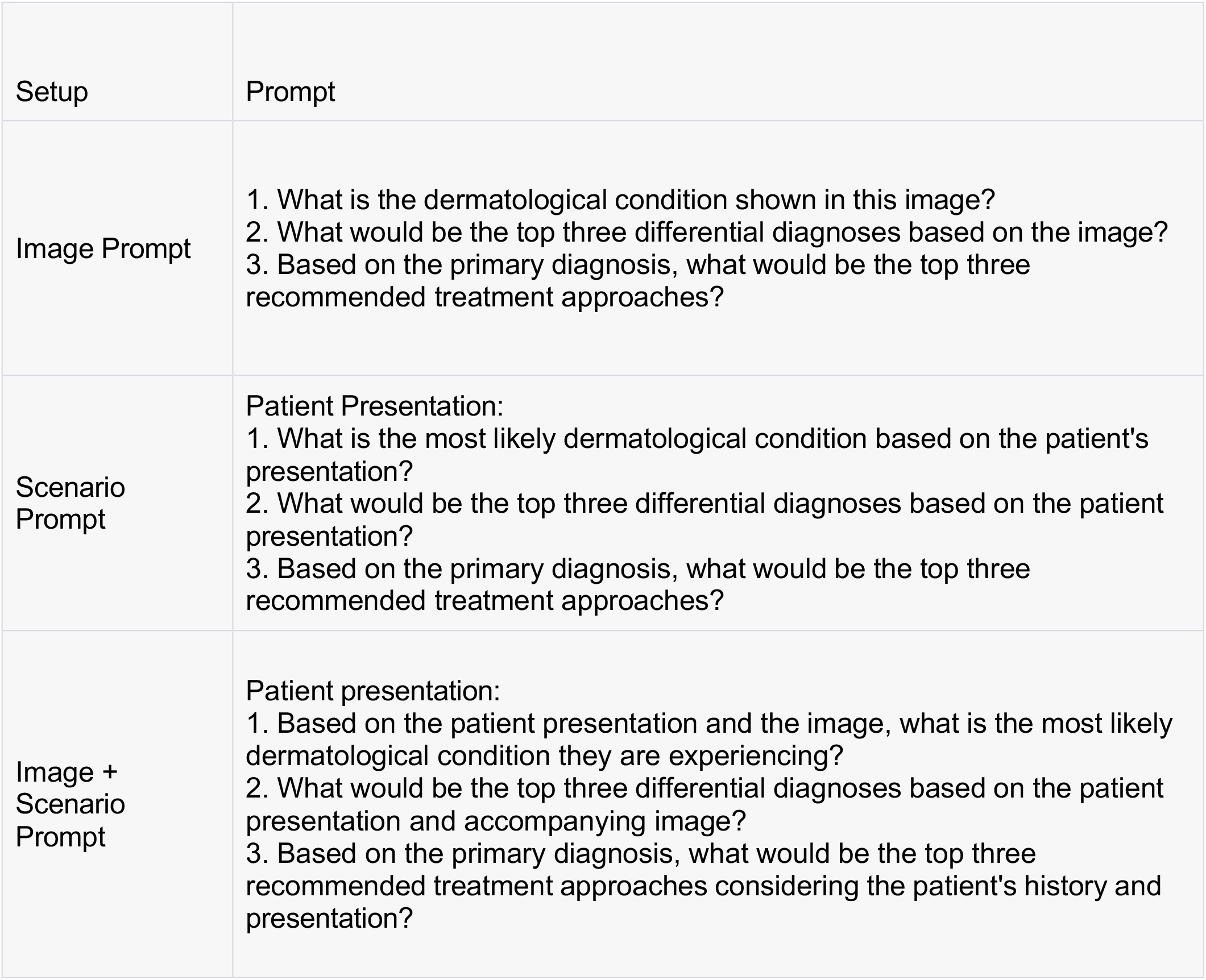
Three distinct setups outlining image prompts, scenario prompts and image and scenario prompts.

| Setup | Prompt |
| --- | --- |
| Image Prompt | 1. What is the dermatological condition shown in this image?<br>2. What would be the top three differential diagnoses based on the image?<br>3. Based on the primary diagnosis, what would be the top three recommended treatment approaches? |
| Scenario Prompt | Patient Presentation:<br>1. What is the most likely dermatological condition based on the patient's presentation?<br>2. What would be the top three differential diagnoses based on the patient presentation?<br>3. Based on the primary diagnosis, what would be the top three recommended treatment approaches? |
| Image + Scenario Prompt | Patient presentation:<br>1. Based on the patient presentation and the image, what is the most likely dermatological condition they are experiencing?<br>2. What would be the top three differential diagnoses based on the patient presentation and accompanying image?<br>3. Based on the primary diagnosis, what would be the top three recommended treatment approaches considering the patient's history and presentation? |

**Figure 1:**
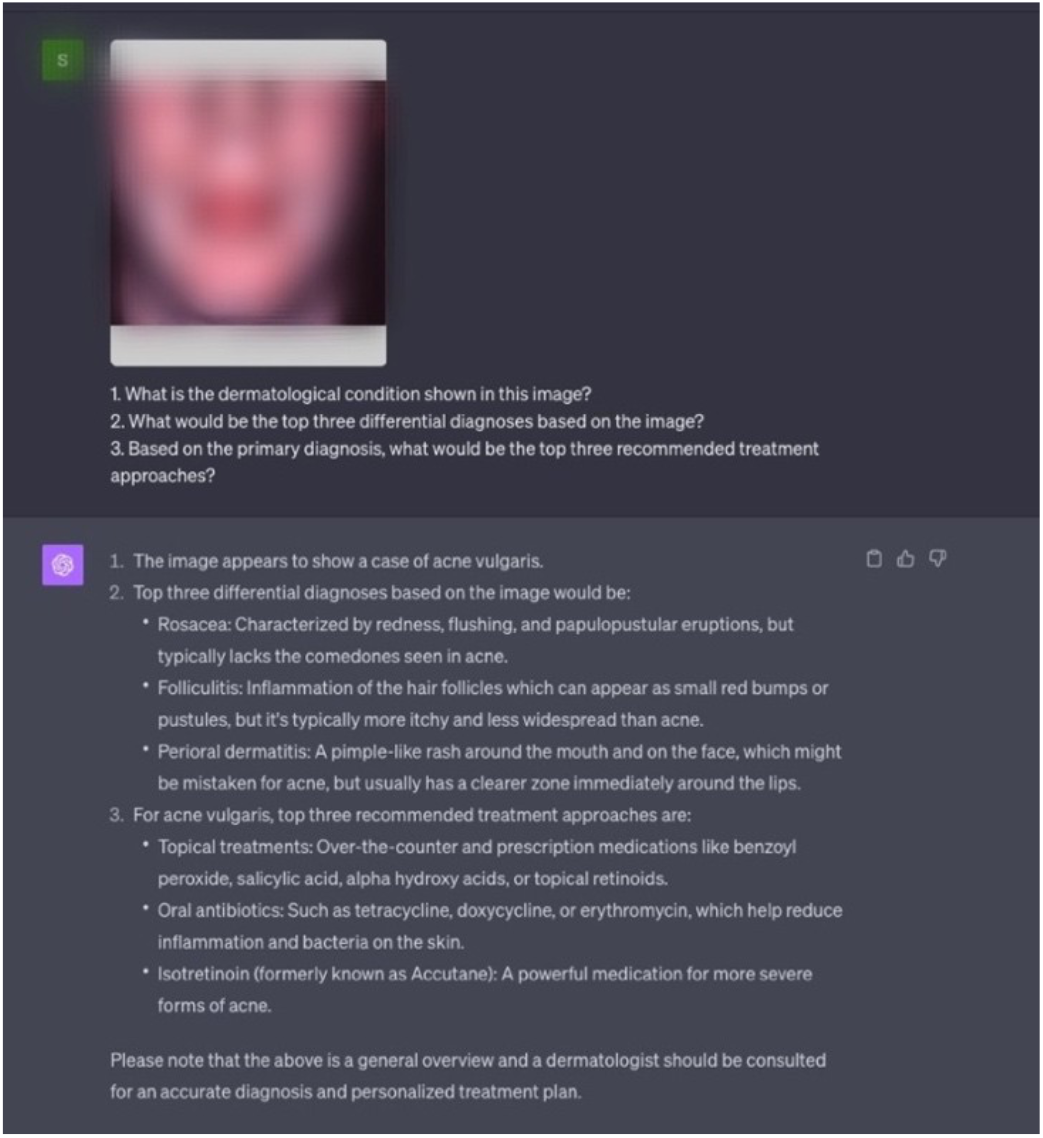
Example of the GPT-4V interface with the image prompt setup. Image blurred out to respect copyright.

In the Image Prompt setup, GPT-4V was provided with images depicting the dermatological condition, and the prompts sought to evaluate its ability to identify the condition, propose differential diagnoses, and suggest treatment approaches solely based on visual input (Figure 1).

In the Scenario Prompt setup, textual descriptions of patient presentations were provided (without a clinical image), and the model was evaluated on its ability to diagnose conditions and suggest treatment approaches based on text input alone.

The Image and Scenario Prompt setup involved a multimodal input scenario where GPT-4V received both image data and textual descriptions of patient presentations. For this setup, each presentation was tested alongside a randomly selected image from the set corresponding to the same diagnosis. This aimed to evaluate the model’s ability to synthesize information from both text and image data to provide accurate diagnoses and treatment recommendations.

### Data Analysis

The analysis was structured to evaluate the diagnostic and treatment recommendation performance of GPT-4V across the three distinct setups. The evaluation metrics were tailored to each type of task—diagnosis, differential diagnosis, and treatment recommendation.

#### Diagnostic Accuracy

The accuracy in diagnosis was assessed by comparing the model’s responses against the established ground truth. For the Image Prompt, the ground truth was ascertained from the database from which the images were sourced, alongside the verification by two board-certified dermatologists. For the Scenario prompt, the ground truth was established based on the dermatologist verification of the scenarios. The Image + Scenario section utilized the same verification methods.

#### Differential Diagnosis

The model’s efficacy in providing a differential diagnosis was evaluated by checking if the correct diagnosis was listed within the top three differential diagnoses generated by the model. This evaluation was consistent across all setups to assess the model’s ability to encompass a range of plausible conditions based on the provided input.

#### Treatment Recommendation

The appropriateness of the model’s treatment recommendations for the Image + Scenario analysis and case-alone analysis was assessed by two dermatologists (JH and DT) using a modified version of an Entrustment Scale sourced from McMaster University’s postgraduate medical education “Competency by Design” evaluation model, which has been adopted by the Royal College of Canada for residency education^8^. Each correctly diagnosed scenario, and image + scenario treatment outcome was rated on a scale from 1-5: 1 -Model did not do task or needed complete guidance, 2 -Model required constant direction, 3 -Model had some independence but required intermittent direction, 4 -Independent but required supervision for safe practice and may have missed some nuances, 5 -Complete independence/understands risks/practice ready. The dermatologists independently rated the relevance and appropriateness of the treatment recommendations provided by the model for each condition. The evaluations from both dermatologists were then averaged to derive a consensus score and a t-test was performed to assess for any differences in performance in the image + scenario group versus the case-only group.

## Results

### Image-Only Analysis

In the image-only analysis, out of the 54 images, the correct primary diagnosis was provided for 29 images (Table 2). Rosacea had the highest diagnostic accuracy, where the correct primary diagnosis was provided for 6/6 of the images. Eczema and squamous cell carcinoma had the lowest diagnostic accuracy, where the correct primary diagnosis was provided for 1/6 images within each condition (Table 3). When asked for the primary diagnosis, the model provided an answer for all 54 images with exception of one image in the superficial spreading melanoma group, where it described the characteristics of the lesion but failed to provide a diagnosis.

**Table 2:** Diagnosis, differential and treatment results for three distinct setups.

| Assessment | Correct primary diagnosis: No. (%) | Correct diagnosis identified in the differential: No. (%) | Average treatment Entrustment Score (0-5) |
| --- | --- | --- | --- |
| Image (n=54) | 29 (54) | 27 (50) | N/A |
| Scenario (n=9) | 8 (89) | 5 (56) | 3.933 |
| Both (n=9) | 8 (89) | 4 (44) | 4.067 |

**Table 3:** Image-only diagnosis and differential diagnosis performance by condition.

| Conditions | Acne | Psoriasis | BCC | Eczema | AK | Rosacea | SCC | SSM | Vitiligo | Total |
| --- | --- | --- | --- | --- | --- | --- | --- | --- | --- | --- |
| Correct diagnosis provided in question 1: | 5/6 | 4/6 | 4/6 | 1/6 | 2/6 | 6/6 | 1/6 | 4/6 | 3/6 | 29/54 |
| Correct diagnosis in the differential | 2/6 | 2/6 | 5/6 | 4/6 | 1/6 | 3/6 | 4/6 | 5/6 | 1/6 | 27/54 |
| Correct Primary diagnosis OR differential | 5/6 | 5/6 | 6/6 | 4/6 | 2/6 | 6/6 | 4/6 | 5/6 | 3/6 | 39/54 |

When assessing diagnostic accuracy with image-only inputs, GPT4V correctly identified the primary condition in 54% of the cases and included the correct diagnosis in the differential list in 50% of the cases (Table 2).

### Scenario-Only Analysis

When provided with text-based scenarios, the model achieved 89% accuracy in identifying the primary diagnosis. The correct diagnosis was included in the list of differential diagnoses in 56% of cases (Table 2). The model provided an incorrect primary diagnosis for only one of nine cases (squamous cell carcinoma) in the scenario prompt, and this was rectified in the list of differential considerations.

### Image + Scenario Analysis

The multimodal analysis, which combines both an image and clinical scenario, resulted in an 89% accuracy rate in identifying the primary diagnosis. The correct diagnosis was provided in the list of differential diagnoses in 44% of the cases (Table 2). The model provided an incorrect primary diagnosis for only one of nine cases (squamous cell carcinoma) in the image and clinical scenario prompt and the differential diagnosis did not capture the correct condition.

Considering the comparable performance of GPT4V in the scenario-only analysis and the multimodal analysis, further testing was conducted to determine whether the model is influenced by one modality over the other. The model was further evaluated on two unique cases to assess image and text preference:

1. A correctly diagnosed image presented with an incorrectly diagnosed clinical scenario.
2. Two incorrectly diagnosed images presented with their correctly diagnosed clinical scenarios.

In the first case, an accurately diagnosed image of squamous cell carcinoma was combined with its incorrectly diagnosed clinical scenario. The results showed an incorrect primary diagnosis of the condition, without rectification in the list of top three differential diagnoses.

In the second case, we evaluated the data from our previous multimodal input analysis and discovered two cases where an incorrectly diagnosed image, both in terms of primary and differential diagnoses, were presented with their correctly diagnosed clinical scenarios. This yielded a correct diagnosis of the primary condition in both cases. The list of differential diagnoses did not include the correct diagnosis.

### Entrustment Score Results

In the assessment of treatment recommendations, the average Entrustment Score was higher in the image and scenario group compared to the scenario only group, however a one-tailed paired-t-test was performed, and it showed no significant difference in performance by the model in providing treatment outcomes (0.14%, 95% CI −0.32% to 0.60%; *P* =.27) (Table 2). Furthermore, the interrater agreeability for this assessment was 37.5%, indicating a moderate level of agreement between the raters involved in evaluating the treatment outcomes.

## Discussion

### Diagnostic Accuracy with Images

In the realm of AI-driven diagnostics, the performance of GPT-4V in dermatology offers a multifaceted view into the current capabilities and limitations of advanced language models in medical applications. The observed diagnostic accuracy rate of 54% from image-only inputs was underwhelming, especially given the extensive pre-training of GPT-4V on a vast corpus of internet images. However, the model card explicitly reveals a lack of specialized training on medical images^1^. A strategic approach of aligning and fine-tuning the model on domain-specific datasets could potentially bridge this accuracy gap. For instance, the Med-PaLM M model, Google’s generalist multimodal medical model, was developed by fine-tuning and aligning the PaLM-E model to the biomedical domain^9^. This process significantly enhanced its performance, achieving a Macro-AUC of 97.27% on PAD-UFES-20, a dermatology dataset. The exemplary performance of Med-PaLM M accentuates the potential for boosting diagnostic accuracy by aligning GPT-4V with the medical domain.

The high diagnostic accuracy for rosacea (6/6) juxtaposed with the low accuracy for eczema (1/6) and SCC (1/6) illustrates a pronounced variability in image-based diagnostic performance across different conditions (Table 3). Moreover, OpenAI’s researchers pinpointed inconsistencies in medical imaging interpretation, underscoring a need for further alignment to improve medical diagnostic accuracy^1^.

### Differential Diagnosis Considerations

The differential diagnosis showed the correct primary diagnosis 50% of the time, despite a primary diagnosis accuracy of 53.7% (Table 1). The ranking of the suspected dermatological conditions indicates a potential misalignment within the model’s hierarchical reasoning (Supplementary table 2). Ensuring that the model accurately reflects the diagnostic significance attributed by medical professionals to the leading diagnosis is crucial for enhancing overall model reliability.

### Text vs Image Priority

The analysis revealed a discernible tendency of the model to prioritize text over visuals, especially in cases where incorrect image diagnoses were corrected by textual diagnoses (Image and Scenario Analysis). This tendency was further exemplified in the case where a correctly diagnosed image, when coupled with an incorrectly diagnosed clinical scenario, led to an overall incorrect diagnosis. The striking disparity in the model’s proficiency in text-only scenarios (89%) compared to image-only scenarios (54%) (refer to Table 2) accentuates a discernible bias towards textual data.

### Performance on Straightforward Cases

The disparity between the 89% diagnostic accuracy on text scenarios tailored for medical students and the sub-60% accuracy on ostensibly simpler image-based cases hints at foundational knowledge gaps (Table 2). Additional training on basic diagnostic examples could potentially ameliorate this issue, better aligning the model’s performance with diagnostic expectations.

### Linking Diagnosis and Treatment

The model demonstrated a logical aptitude in recommending treatment options yet faltered in connecting image details to treatment nuances, further underscored by the lack of a significant difference in the Dermatologist ratings between the scenario-only and the image and scenario setups (Table 1, Average Treatment Entrustment Score). Feedback from one dermatologist likened the performance of GPT-4V to an early dermatology resident or an interested medical student, suggesting that the unaligned generalist model does have some promise to work alongside a doctor to assist, rather than TO diagnose and treat. Augmenting the contextualization of images in relation to personalized therapeutic plans could significantly enhance the model’s clinical applicability.

Another dermatologist compared GPT-4’s treatment capabilities to those of a fourth-year medical resident, highlighting its proficiency in textbook treatments for various conditions. However, real-life clinical practice encompasses additional layers of decision-making that are crucial. For instance, in treating melanoma, GPT-4V tends to suggest advanced treatment options prematurely, overlooking essential steps like initial biopsy, formal diagnosis, and staging (Supplementary Table 3). These steps are vital in actual clinical scenarios. Similarly, while its approach to vitiligo treatment is technically accurate, it fails to account for the contextual and geographical nuances that significantly influence treatment decisions in real-life cases. These insights emphasize the important role of physicians in providing comprehensive care and highlight the necessity for enhanced contextual awareness in medical AI applications, extending beyond just textbook accuracy.

### Entrustability Scale Limitations

The scale used to assess these treatment outcomes, despite offering a structured approach, has its limitations. The inherent subjectivity of the scale and its broad descriptors can lead to variability in evaluations, indicated by the moderate inter-rater agreeability score of 37.5%. Additionally, this scale emphasizes the model’s independence in decision-making but may not accurately reflect the effectiveness or appropriateness of the proposed treatments. The risk of overestimating the AI’s capabilities through this scale suggests a need for more nuanced and context-specific assessment methods to appropriately evaluate such models in future studies.

### Limitations and Inherent Biases of AI in Dermatology

The integration of Large Language Models (LLMs) into dermatology offers notable benefits for medical education and can enhance clinical efficiency as supplementary tools. However, their application in healthcare settings risks reinforcing existing biases and racial disparities. Dermatology’s historical tendency to focus on white skin tones in research and educational resources has led to a significant underrepresentation of richly pigmented skin^10,11^. Consequently, AI models, often trained on datasets dominated by images of patients with white skin, may inadvertently perpetuate these disparities. A 2018 study demonstrated the superior performance of deep-learning convolutional neural networks in identifying potentially cancerous skin lesions compared to most dermatologists^12^. However, the study’s limitation was its predominant use of images sourced from the International Skin Imaging Collaboration: Melanoma Project, which primarily complied data from dermatological conditions on white skin from populations in the USA, Europe, and Australia^13^. This bias in AI training can result in less accurate diagnoses or delayed recognition of skin conditions in patients with darker skin tones, hindering efforts to achieve equitable healthcare.

As LLMs become more prevalent in the medical field, their potential to impact quality of care grows^14^. This underscores the urgent need for comprehensive regulatory frameworks. Such frameworks should mandate rigorous testing of these AI tools to identify and mitigate biases across different skin types, ensuring they adhere to stringent standards of accuracy and fairness.

Moreover, dermatologists and healthcare professionals employing these AI tools must be cognizant of their inherent limitations and biases. It is imperative that when using AI tools, they maintain a high level of clinical judgment, particularly when treating patients from diverse racial backgrounds. This approach will help ensure that the deployment of LLMs in dermatology contributes positively to patient care, respecting the nuances of diverse skin types and promoting equitable healthcare practices.

## Conclusion

This analysis sheds light on the limitations inherent in generalist pre-training and pinpoints avenues for improvement in hierarchical reasoning, multimodal integration, foundational medical knowledge, and contextual reasoning for clinical AI. It also accentuates the significance of fine-tuning and medical collaboration in unlocking AI’s potential to augment dermatological practice. The insights provided by dermatologists, likening GPT-4’s performance to that of a dermatology resident reiterate the necessity for refined training and evaluation to ensure safety and efficacy in clinical applications.

## Data Availability Statement

The images used in this study can be accessed on dermnet.nz and dermatlas.org. Additional data collected for this study is included in the supplementary materials. This includes the case scenarios created for each dermatological condition, the differential diagnosis breakdown for the image-only analysis, and the top 3 treatment recommendations per scenario in the combined image and scenario analysis.

## Abbreviations

AI: Artificial Intelligence
LLM: Large Language Model
GPT: Generative Pre-trained Transformer

