## Supplemental Materials for "Evaluating the Diagnostic and Treatment Recommendation Capabilities of GPT-4 Vision in Dermatology"

### SUPPLEMENTARY MATERIALS

Supplementary Table 1: Case Scenarios for nine common dermatological conditions.

| |
|---|
| 1. Acne: |
| A 13-year-old boy presents to the family clinic with no significant past medical history. He plays numerous sports at school such as wrestling, and football. On physical exam, you observe numerous lesions on his face, along with his chest, and back. |
| 2. Eczema (Atopic Dermatitis) |
| A mother presents to the family clinic concerned about her 5-month-old baby. She shares that her baby has developed redness around his face, especially around the cheeks and chin, as well as on his chest and elbows. She is concerned that her baby's skin is very dry and irritated. |
| 3. Psoriasis |
| A 65-year-old female presents to your family medicine clinic with back pain that she has been experiencing for the past year. She describes morning stiffness that lasts about an hour upon waking and states. She states that another reason for her visit is it address concerns regarding her skin. She rolls up her sleeves and shares that she has also been observing patches of dry, scaly skin on her elbows and sometimes on her knees. "This skin thing must be getting bad because even my nails look different according to my nail-tech." |
| 4. Actinic Keratoses |
| A 65-year-old white female presents with multiple pink lesions on her temples and forehead. She initially thought that these scaly spots may have been a sign of dryness, but they have persisted for months and are rough and bumpy to the touch. She has no past medical history other than a diagnosis of hypertension which is well managed with medications. |
| 5. Rosacea: |
| A 40-year-old male presents with facial flushing. He states that this has been fluctuating for decades, but now the redness has become more persistent. When asked about provoking factors, he says that the redness is worse after he goes on walks in the afternoon, practices yoga or when he drinks alcohol. He notes that in the past he has experienced stinging sensations in the same areas where the redness occurs. |
| 6. BCC |
| A 68-year-old white male presents with a lesion on the scalp. His wife was the first to notice it and she thought it might have been a shiny pimple or bug bite, but it has persisted for months and appears to be growing. Patient describes no pain to the touch. He has no past medical history of note. |
| 7. SCC |
| A 75-year-old white female presents with concerns about a lesion on her chest. She was previously treated with localized radiotherapy for lung cancer at the site of the lesion of |

| |
|---|
| concern. She is worried about the appearance of the lesion as it “looks like an open sore,” and she finds herself itching the area which often causes bleeding. |
| 8. Superficial Spreading Melanoma |
| A 42-year-old white male presents with a black and dark-brown lesion on his back. He states that his partner had noticed something new there 9 months ago when they went on vacation. Over the past few months, he has been growing increasingly worried because it is getting bigger. |
| 9. Vitiligo |
| A 29-year-old black female presents with white patches on her skin. She states that she first started to note small patches of skin without pigment on her hands, but that over the course of a few months they are getting bigger and showing up in other places, such as her face and feet. She has a past medical history of type 1 diabetes that is treated to target and no other medical conditions. |

Supplementary Table 2: Image-only prompt performance across nine dermatological conditions and differential ranking.

| Conditions | Acne | Psoriasis | BCC | Eczema | AK | Rosacea | SCC | SCM | Vitiligo | Total |
|---|---|---|---|---|---|---|---|---|---|---|
| Correct diagnosis provided in question 1: | 5/6 | 4/6 | 4/6 | 1/6 | 2/6 | 6/6 | 1/6 | 4/6 | 3/6 | 29/54 |
| No diagnosis provided in question 1: | 0/6 | 0/6 | 0/6 | 0/6 | 0/6 | 0/6 | 0/6 | 1/6 | 0/6 | 1/54 |
| Correct diagnosis provided #1 in the differential | 0/6 | 1/6 | 2/6 | 2/6 | 0/6 | 0/6 | 0/6 | 1/6 | 0/6 | 6/54 |
| Correct diagnosis provided #2 in the differential | 0/6 | 0/6 | 0/6 | 1/6 | 0/6 | 0/6 | 2/6 | 0/6 | 0/6 | 3/54 |
| Correct diagnosis provided #3 in the differential | 0/6 | 0/6 | 0/6 | 0/6 | 0/6 | 0/6 | 1/6 | 0/6 | 0/6 | 1/54 |
| Correct Primary diagnosis/considered in differential | 5/6 | 5/6 | 6/6 | 4/6 | 2/6 | 6/6 | 4/6 | 5/6 | 3/6 | 40/54 |

Supplementary table 3: Image + scenario prompt performance with corresponding treatment recommendations.

| Condition | Diagnosis | Differential | Treatment |
|---|---|---|---|
| Acne | Correct | 1. Acne Vulgaris<br>2. Folliculitis<br>3. Rosacea | 1. Topical retinoids<br>2. Benzoyl peroxide preparations<br>3. Oral antibiotics |
| Psoriasis | Correct | 1. Psoriatic arthritis,<br>2. Atopic dermatitis,<br>3. Lichen Planus | 1. Topical treatments<br>2. Light therapy (phototherapy)<br>3. Systemic medications |
| BCC | Correct | 1. SCC<br>2. Actinic Keratosis<br>3. Seborrheic Keratosis | 1. Surgical Excision<br>2. Mohs Micrographic Surgery<br>3. Topical treatments |
| Eczema | Correct | 1. Seborrheic dermatitis<br>2. Contact dermatitis.<br>3. Psoriasis | 1. Moisturizing<br>2. Topical corticosteroids<br>3. Avoiding triggers |
| Actinic Keratosis | Correct | 1. Seborrheic Keratosis<br>2. BCC<br>3. SCC | 1. Topical Chemotherapy<br>2. Cryotherapy<br>3. Photodynamic Therapy |
| Rosacea | Correct | 1. Rosacea<br>2. Seborrheic Dermatitis<br>3. Photodermatitis | 1. Topical Medications<br>2. Oral Antibiotics<br>3. Laser and Light Therapy |
| SCC | Incorrect | 1. Chronic radiation dermatitis<br>2. Basal cell carcinoma<br>3. Actinic keratosis | 1. Topical corticosteroids<br>2. Emollients<br>3. Wound care |
| Superficial Spreading Melanoma | Correct | 1. Melanoma<br>2. Dysplastic nevus<br>3. Seborrheic keratosis | 1. Surgical excision<br>2. Sentinel lymph node biopsy<br>3. Adjuvant therapy |
| Vitiligo | Correct | 1. Vitiligo<br>2. Pityriasis Alba<br>3. Tinea Versicolor | 1. Topical Therapy: topical Steroids, topical Calcineurin Inhibitors<br>2. Phototherapy: narrowband UVB therapy<br>3. Surgical Options: skin grafting, melanocyte transplantation. |
